# SARS-CoV-2 naïve and recovered individuals show qualitatively different antibody responses following mRNA vaccination

**DOI:** 10.1101/2021.05.07.21256821

**Authors:** Sonia Tejedor Vaquero, Leire de Campos-Mata, José María Ramada, Pilar Díaz, Juan Navarro-Barriuso, Clara Ribas-Llaurado, Natalia Rodrigo Melero, Carlo Carolis, Andrea Cerutti, Ramon Gimeno, Giuliana Magri

## Abstract

mRNA-based vaccines effectively induce protective neutralizing antibody responses against SARS-CoV-2, the etiological agent of COVID-19. The specific compositional patterns of these responses remain largely unknown. We found that SARS-CoV-2-naïve individuals receiving the first dose of an mRNA vaccine developed a SARS-CoV-2-specific antibody response with a subclass profile comparable to that induced by the natural infection, except IgA2, which did not increase. SARS-CoV-2-naïve subjects also mounted a robust virus-specific recall response after receiving the second dose. This response increased all IgG subclasses, but boosted neither IgM nor IgA1 and IgA2 subclasses. In contrast, individuals recovered from COVID-19 mounted peak virus-specific antibody responses upon primary immunization and did not further augment such responses following secondary immunization. Remarkably, compared to SARS-CoV-2-naïve subjects, individuals with pre-existing immunity showed increased levels of all virus-specific antibodies but IgG3 following primary vaccination. By dissecting the heterogeneity of mRNA vaccine-induced humoral responses to SARS-CoV-2, our findings indicate that the induction of optimal immune protection may require the development of personalized vaccination programs.

## Introduction

Vaccination against Severe Acute Respiratory Syndrome Coronavirus 2 (SARS-CoV-2) is the leading option to achieve protection against the coronavirus disease 19 (COVID-19) pandemic^1^. SARS-CoV-2 messenger RNA (mRNA)-based vaccines, such as the “mRNA-1273” from Moderna or the “BNT162b2” from Pfizer-BioNTech have been reported to be safe and highly effective in preventing severe COVID-19 disease in clinical trials^2,3^. Hence, they have been recently approved for emergency use and are currently being administrated to millions of individuals in numerous Western countries. BNT162b2 and mRNA-1273 vaccines encode for the full-length spike (S) protein of SARS-CoV-2, which is essential for viral pathogenicity, and are applied in a two-dose immunization regimen, administered 21 days or 28 days apart, respectively.

Earlier phase III trials and more recent studies have demonstrated that both mRNA-based vaccines induce a strong anti-S IgG humoral response and promote the generation of S-specific memory T and B cells after the two-dose regimen^2–8^. Among the different antibodies induced upon vaccination, the ones that specifically recognize the receptor binding domain (RBD) of the viral S protein are particularly relevant as they mirror the neutralizing capacity of the sera^9,10^ and are considered a principal surrogate of immune protection and vaccine efficacy.

Interestingly, recent studies have shown that SARS-CoV-2 naïve individuals and individuals with prior history of infection have distinct immune responses upon mRNA vaccination. Following the first vaccine dose, COVID-19 recovered individuals show significantly higher S-and RBD-specific IgG titers, superior serum neutralization activity^7,11–13^, even against SARS-CoV-2 variants^8,14,15^, and increased S protein-specific memory T and B cell responses than naïve individuals^4–6,14^. This enhanced response is consistent with the persistence of humoral and cellular immunity against SARS-CoV-2 in convalescent individuals, as previously reported^16–20^. Because a recall immunization does not seem to provide any quantitative benefit in subjects with pre-existing immunity to SARS-CoV-2^6,7,13^, it has been proposed that COVID-19 recovered people may only require a single vaccine dose to achieve optimal immune protection^6,7,13^.

These studies have highlighted the importance of understanding the immunological history of an individual to evaluate the efficacy of vaccines against SARS-CoV-2. However, to date, the analysis of humoral responses to SARS-CoV-2 vaccines has exclusively focused on measuring S protein-specific total IgG responses. The contribution of virus-specific IgG subclasses and antibody classes different from IgG to vaccine-induced humoral responses remains elusive.

In humans, IgG consists of IgG1, IgG2, IgG3 and IgG4 subclasses, which differ in terms of structure, effector functions and serum half-life^21^. IgG1 and IgG3 are the most abundant antibodies produced upon viral infection and are required for anti-viral immunity. These IgG subclasses activate the complement cascade, mediate antibody-dependent cellular cytotoxicity, and stimulate phagocytic cells via powerful receptors termed Fcγ receptor I (FcγRI) and FcγRIII. In contrast, IgG2 and IgG4 subclasses neither activate complement nor signal through FcγRI and FcγRIII and are poorly induced upon viral infection^22^.

Recent studies, including ours^10,16,23^, have shown that IgG1 and IgG3 subclasses are indeed strongly induced soon after SARS-CoV-2 infection, whereas IgG2 and IgG4 subclasses are induced to a much lesser extent. In recovered individuals, humoral and B cell memory responses appeared to be IgG1-dominated^16,19^. SARS-CoV-2 infection also induces virus-specific IgM, IgA1 and, to a lesser extent, IgA2, which all wane during convalescence^16^.

How RBD-specific IgG and IgA subclasses are differentially induced upon administration of mRNA vaccines in SARS-CoV-2 naïve and recovered individuals is unknown. The characterization of vaccine-induced RBD-specific antibody profiles with subclass resolution may enhance our knowledge of humoral correlates of protection.

Here we characterized serum RBD-specific antibody classes and subclasses, including IgG1, IgG2, IgG3, IgG4, IgM, IgA1 and IgA2 in a longitudinal cohort of SARS-CoV-2 naïve and COVID-19 recovered individuals who received and mRNA-based vaccine. Our analysis revealed striking differences in the quantity and quality of antibody responses to the vaccine based on the prior history of infection. Our findings may offer new insights into the development of personalized vaccine strategies for the induction of optimal immune protection.

## Results

For this study, we recruited 48 healthy individuals who received SARS-CoV-2 mRNA vaccines (Moderna mRNA-1273 or Pfizer-BioNTech BNT162b2) at Parc Salut Mar (Barcelona, Spain). Of this cohort, 20 individuals had previous SARS-CoV-2 infection (mean age = 41; 75% female) and 28 were SARS-CoV-2 naïve (mean age = 37, 68% female) (**Fig. 1A**). Sera were collected at three time points: pre-vaccine baseline (B), 2/3 weeks post the first dose (PFD), and one month post the second dose (PSD) (**Fig. 1B**).

**Fig. 1.**
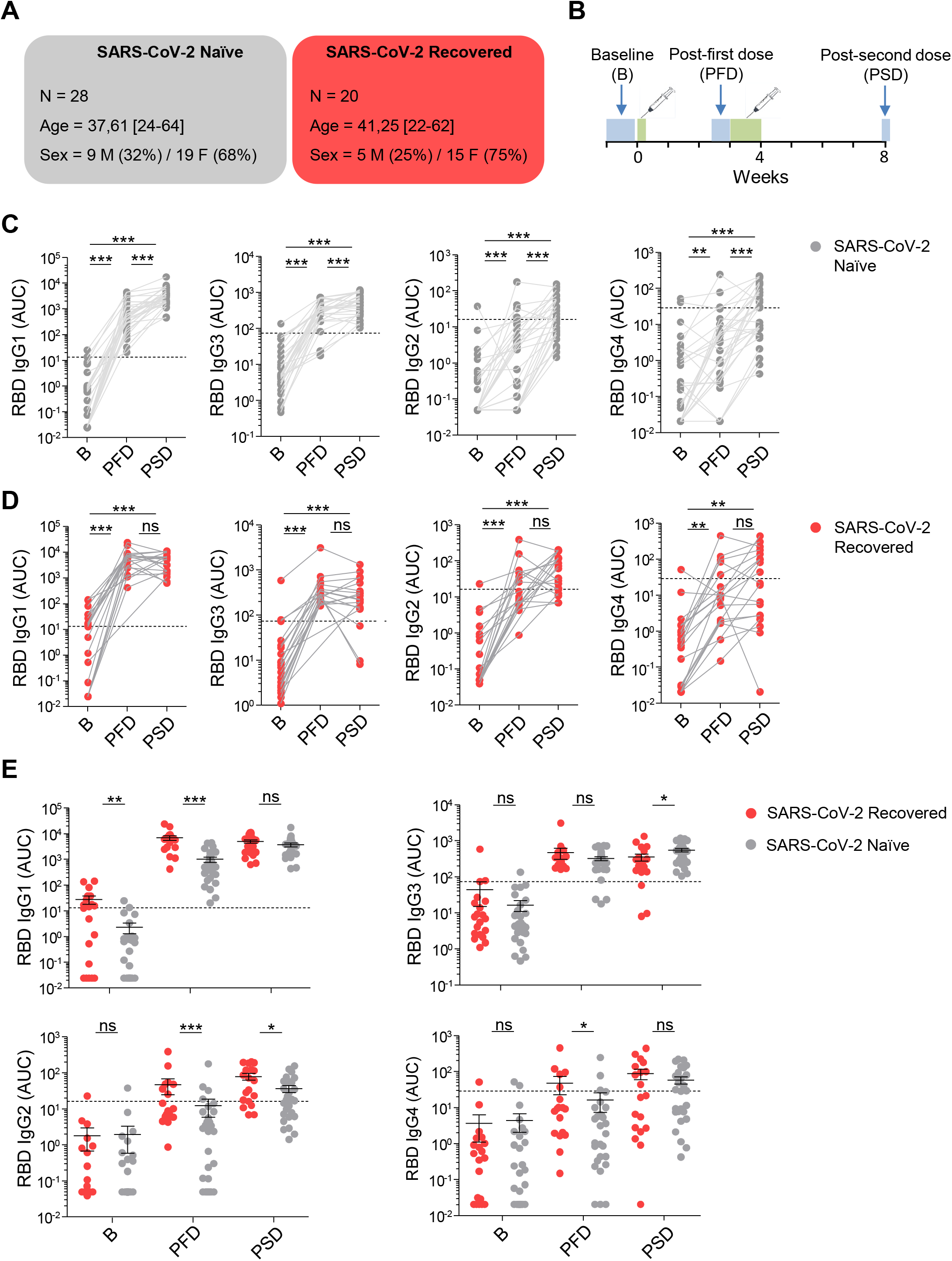
mRNA vaccination induces different pattern of RBD-specific IgG subclass responses in SARS-CoV-2 naïve and recovered individuals. (**A**) Schematic diagram of the cohort characteristics. (**B**) Schematic diagram of the study timeline. Sera were collected at 3 time points: pre-vaccine baseline (B), 2/3 weeks following first dose (PFD) and 1 month following the second dose (PSD). Range of days per time point is indicated with a box. (**C**) Area under the curve (AUC) for each of the RBD-specific IgG subclasses analyzed from SARS-CoV-2 naïve individuals overtime. (**D**) AUC for each of the RBD-specific IgG subclasses analyzed from SARS-CoV-2 recovered individuals overtime. (**E**) AUC for each of the RBD-specific IgG subclasses analyzed from SARS-CoV-2 naïve and recovered individuals overtime depicted in the same graph. Sera from SARS-CoV-2 naïve individuals at baseline were used to establish negative threshold values defined as the naïve AUC mean plus 2 times the standard deviation of the mean. Dashed line indicates negative threshold. Data are presented as individual dots. Two-tailed Mann-Whitney U test was performed to compare time points. Wilcoxon matched pairs test was performed to compare SARS-CoV-2 naïve and recovered individuals (^*^P < 0.05, ^**^P < 0.01, and ^***^P < 0.001). SARS-CoV-2 Naïve, n=28; SARS CoV-2 recovered, n=20.

To elucidate whether SARS-CoV-2 naïve and recovered individuals can mount comparable humoral responses to mRNA vaccines, we used a previously described^23,24^ two-step enzyme-linked immunosorbent assay (ELISA) that determines antibody responses to the RBD of the S protein from SARS-CoV-2. To quantitate antibodies, we performed serial dilutions of the serum samples and used the optical density (OD) values to determine the area under the curve (AUC).

First, we evaluated the induction of RBD-specific IgG subclasses upon mRNA vaccination in SARS-CoV-2 naïve individuals and individuals with prior history of SARS-CoV-2 infection. The administration of the first dose of the mRNA vaccine in naïve individuals induced a significant increase of all RBD-specific IgG subclasses, which further increased following the second dose. As expected, the highest increase was observed for RBD-specific IgG1 and to a lesser extent, RBD-specific IgG3, whereas RBD-specific IgG2 and IgG4 showed much lower increases (**Fig. 1C**). In subjects with pre-existing immunity, we also observed a significant increase of all RBD-specific IgG subclasses after the first dose of the mRNA vaccine; however, in these individuals the booster dose did not induce a further increase of any IgG subclasses (**Fig. 1D**).

We then compared the levels of anti-RBD IgG1, IgG3, IgG2 and IgG4 at baseline, after the first dose, and one month after the second dose between SARS-CoV-2 naïve and recovered individuals (**Fig. 1 E**). At baseline, study participants with prior history of infection showed significantly higher levels of RBD-specific IgG1 compared to SARS-CoV-2 naïve individuals. In contrast, no significant differences were observed between these two groups with respect to other IgG subclasses. After the first dose of an mRNA vaccine, RBD-specific IgG1, IgG2 and IgG4 titers in recovered individuals were significantly higher than in vaccinees with no pre-existing immunity to SARS-CoV-2. Interestingly, after primary immunization, the amount of RBD-specific IgG3 was similar in SARS-CoV-2 naïve and COVID-19 recovered individuals. Following the second dose of the vaccine, the concentrations of RBD-specific IgG1 and IgG4 were similar between the two groups of vaccinees, whereas RBD-specific IgG2 levels were still more elevated in recovered individuals compared to SARS-CoV-2 naïve subjects. Remarkably, one month after the reboost, individuals with no prior history of SARS-CoV-2 infection showed slightly more RBD-specific IgG3 compared to recovered individuals (**Fig. 1E**). Of note, we did not observe any relationship between sex and RBD-specific IgG subclasses at baseline, after the first immunization and after the recall immunization (**Fig. S1A**).

Recent studies reported that mRNA vaccination induced S-specific IgA and IgM along with S-specific IgG in fully vaccinated individuals^12^. Moreover, circulating plasmablasts producing IgA anti-S were detected one week after the second dose of mRNA vaccine^25^. Yet, how S-specific IgM as well as IgA1 and IgA2 subclasses are differentially induced upon vaccination in SARS-CoV-2 naïve and COVID-19 recovered individuals remains unclear. Our analysis revealed that RBD-specific IgM increased after primary vaccination, but it did not further augment upon reboost in both groups of vaccinees (**Fig. 2A, B**). We further evaluated the level of RBD-specific IgA1 and IgA2 subclasses. While IgA1 induction occurs in both mucosal and systemic compartments, IgA2 production is largely confined to the intestinal compartment^26^. Similarly to RBD-specific IgM, RBD-specific IgA1 was induced upon the first dose of vaccine in SARS-CoV-2 naïve and recovered individuals, but its serum concentration no longer augmented one month following the second dose (**Fig. 2A, B**). RBD-specific IgA2 titers were below the threshold at baseline and after the first and second dose of mRNA vaccine in SARS-CoV-2 naïve individuals (**Fig. 2A**), whereas a small but significant induction was observed in individuals with pre-existing immunity after primary but not secondary vaccination (**Fig. 2B**). When we compared the levels of RBD-specific IgM, IgA1 and IgA2 in SARS-CoV-2 naïve and recovered individuals, we observed no significant differences at baseline, being most of AUC values below the threshold. However, after the first and second dose, individuals with prior history of SARS-CoV-2 infection showed significantly higher levels of RBD-specific IgM, IgA1 and IgA2 compared to SARS-CoV-2 naïve individuals (**Fig. 2C**). Again, no association was observed between sex and RBD-specific IgM or IgA subclasses at any time points (**Fig. S1B**).

**Fig. 2.**
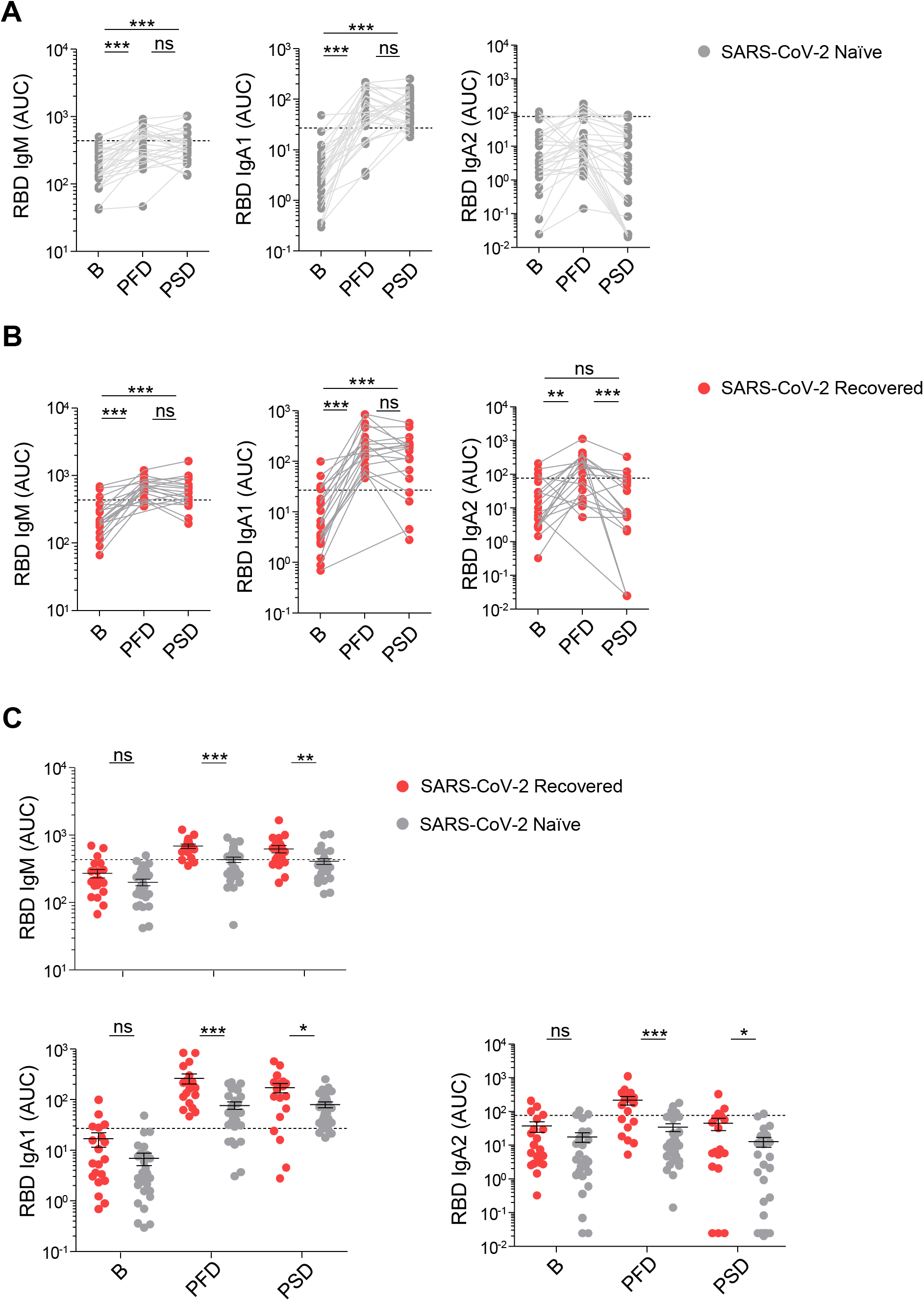
mRNA vaccination induces increased levels of RBD-specific IgM and IgA subclasses in recovered individuals compared to SARS-CoV-2 naive individuals. (**A**) Area under the curve (AUC) for each of the RBD-specific IgM and IgA subclasses analyzed from SARS-CoV-2 naïve individuals overtime. (**B**) AUC for each of the RBD-specific IgM and IgA subclasses analyzed from SARS-CoV-2 recovered individuals overtime. (**C**) AUC for each of the RBD-specific IgM and IgA subclasses analyzed from SARS-CoV-2 naïve and recovered individuals overtime depicted in the same graph. Sera from SARS-CoV-2 naïve individuals at baseline were used to establish negative threshold values defined as the naïve AUC mean plus 2 times the standard deviation of the mean. Dashed line indicates negative threshold. Data are presented as individual dots. Two-tailed Mann-Whitney U test was performed to compare time points. Wilcoxon matched pairs test was performed to compare SARS-CoV-2 naïve and recovered individuals (^*^P < 0.05, ^**^P < 0.01, and ^***^P < 0.001). SARS-CoV-2 Naïve, n=28; SARS CoV-2 recovered, n=20.

## Discussion

Here, we showed that mRNA vaccination induced distinct profiles of subclass-specific antibody responses in SARS-CoV-2 naïve compared to COVID-19 recovered individuals. Our findings highlight both the complexity and heterogeneity of antibody responses to mRNA vaccines against SARS-CoV-2 in individuals with a different exposure history.

Prior to vaccination, only RBD-specific IgG1 was significantly higher in COVID-19 recovered individuals compared to SARS-CoV-2 naïve subjects, which is consistent with the predominance of virus-specific IgG1 and memory B cell responses observed in COVID-19 convalescent individuals^16,19^. Following primary immunization, individuals with pre-existing immunity showed increased titers of all RBD-specific antibodies, except IgG3, when compared to SARS-CoV-2 naïve subjects. This was consistent with the presence of pre-existing SARS-CoV-2-specific class-switched and unswitched memory B cells in COVID-19 recovered subjects^16,17,19,20^. In line with our results, a recent report showed a strong correlation between the frequency of SARS-CoV-2-specific memory B cells at baseline and post-vaccination antibody levels^6^. Moreover, the lack of superior RBD-specific IgG3 responses in COVID-19 recovered individuals is in agreement with earlier findings indicating that very few SARS-CoV-2 memory B cells express IgG3 in COVID-19 convalescent individuals^19^.

Our analysis clearly shows that primary vaccination of SARS-CoV-2 naïve individuals induces a broad spectrum of virus-reactive antibodies responses, which include RBD-specific IgM, IgA1 as well as IgG1, IgG3 and, to a lesser extent, IgG2 and IgG4. Keeping in mind the largely mucosal nature of IgA2^23,26^, it was not surprising to detect virtually no RBD-specific IgA2 induction in individuals with no prior history of infection. Thus, except for the lack of IgA2 induction, the general pattern of subclass-specific antibody responses induced by primary immunization with mRNA vaccines of SARS-CoV-2 naïve individuals was comparable to that seen after natural infection^16^. The small but significant induction of RBD-specific IgA2 in recovered individuals upon the first dose of mRNA vaccine echoes earlier studies that describe IgA2-expressing memory B cells targeting the S protein of SARS-CoV-2 in a subset of COVID-19 recovered patients^27,28^. Remarkably, we previously detected RBD-dominant IgA2 responses mostly in COVID-19 patients experiencing gastrointestinal symptoms^23^. Future studies should evaluate whether there is a relationship between COVID-19 disease-related symptoms and SARS-CoV-2-specific antibody profiles after vaccination in recovered subjects.

Remarkably, differences in vaccine-induced antibody responses among SARS-CoV-2 naïve subjects and COVID-19 recovered individuals were also observed after recall immunization. In contrast to individuals with prior history of infection, SARS-CoV-2 naïve subjects showed a vigorous boosting of RBD-specific IgG responses upon the second dose of mRNA vaccination, which involved all IgG subclasses. Interestingly, after the second injection, RBD-specific IgG2 titers in SARS-CoV-2 naïve individuals were still significantly lower compared to those of recovered subjects, whereas IgG3 titers were significantly higher. The increased levels of RBD-specific IgG3 in SARS-CoV-2 naïve individuals could be due to the induction of increased number of IgG3 expressing memory B cells after primary vaccination or differences in the kinetics of humoral responses. Contrarily, no reboost effect was observed for RBD-specific IgM and IgA subclasses in both vaccinee groups, and the titers of these antibodies were consistently lower in fully vaccinated naïve subjects compared to recovered individuals. The lack of increased antibody response following recall vaccination in individuals with pre-existing immunity to SARS-CoV-2 is intriguing and could relate to the reduced antigen availability resulting from the elevated vaccine-specific IgG responses triggered by the primary immunization. However, it could also be due to differences in the kinetics of germinal center or extra follicular B cell responses to viral antigens as well as differences in the antigen-presenting cell priming. Such differences may influence the generation of secretory memory, including the generation of long-lived plasma cells in the bone marrow. Further studies are required to address these questions. In summary, our findings highlight the heterogeneity of mRNA vaccine-induced humoral responses to SARS-CoV-2. They also indicate that prior viral exposure may be the major determinant of this heterogeneity. Finally, they suggest that the induction of optimal immune protection may require the development of personalized vaccination programs.

## Limitations of the study

This study has the following limitations. First, the size of the studied cohort was small (*n*=48) and it only included SARS-CoV-2 recovered individuals who had experienced mild COVID-19 symptoms. Hence, our results should be confirmed in future larger-scale studies encompassing COVID-19 recovered individuals with more severe disease. In addition, all the participants of our study were below 65 years of age (mean = 39). Older SARS-CoV-2 naïve and COVID-19recovered individuals should be studied in the future to fully address the vaccine efficacy in this specific population. Finally, to fully investigate the protection offered by mRNA vaccines overtime, additional serologic measurements should be performed at later time points.

## Material and Methods

### Contact for Reagent and Resource Sharing

Further information and requests for resources and reagents should be directed to and will be fulfilled by the corresponding author, Giuliana Magri.

### Experimental Model and Subject Details

#### Study Cohort

48 healthy individuals provided written consent and were enrolled in the study. All procedures followed were approved by the Ethical Committee for Clinical Investigation of the Institut Hospital del Mar d’Investigacions Mèdiques (Number 2020/9621/I). Subjects were stratified in two groups: SARS-CoV-2 naïve (*n* = 20) and SARS-CoV-2 recovered (*n* = 28), based on self reported or laboratory evidence of prior SARS-CoV-2 infection. Of the self-reported naïve subjects, one individual was found to have positive SARS-CoV-2 nucleoprotein-specific antibodies at baseline and was retroactively classified as SARS-CoV-2 recovered. All participants classified as SARS-CoV-2 recovered had mild COVID-19 course and symptoms during infection. Mean age were 38 ([24-64]) years old for SARS-CoV-2 naïve group and 41 ([22-62]) years old for SARS-CoV-2 recovered group. From all the participants, 29% were male and 71% were female. Detailed information of groups and patients were summarized in **Fig. 1**. All study participants received either Moderna (mRNA-1273, *n* = 46) or Pfizer (BNT162b2, *n* = 2) mRNA vaccine at Parc Salut Mar (Barcelona, Spain). Blood samples were collected at 3 time points: pre-vaccine baseline (B), 2/3 weeks post first vaccine dose (PFD) and 4 weeks post-second vaccine dose (PSD).

### Production of recombinant SARS-CoV-2 proteins

The pCAGGS RBD construct, encoding for the receptor-binding domain of the SARS-CoV-2 Spike protein (amino acids 319-541 of the Spike protein) along with the signal peptide plus a hexahistidine tag was provided by Dr Krammer (Mount Sinai School of Medicine, NY USA). RBD proteins were expressed in-house in Expi293F human cells (Thermo Fisher Scientific) by transfection of the cells with purified DNA and polyethylenimine (PEI). Cells were harvested 3 days post transfection and RBD-containing supernatants were collected by centrifugation at 13000rpm for 15min. RBD proteins were purified in Hitrap-ni Columns in an automated Fast Protein Liquid Chromatography (FPLC; Äkta avant), concentrated through 10 kDa Amicon centrifugal filter units (EMD Millipore) and resuspended in phosphate buffered saline (PBS).

### Enzyme-linked immunosorbent assay (ELISA)

Sera were collected from whole blood in silica-treated tubes where the blood was incubated for 30 min without movement to trigger coagulation. Next, samples were centrifuged for 10 min at 1300 g at room temperature (RT), heat-inactivated at 56ºC for 1 hour and stored at −20ºC prior to use. ELISAs performed in this study were adapted from previously established protocols^23,24^. 96-well half-area flat bottom high-bind microplates (Corning) were coated overnight at 4ºC with SARS-CoV-2 RBD recombinant viral protein at 2 µg/ml in PBS (30 µl per well). Plates were washed with PBS 0.05% Tween 20 (PBS-T) and blocked with blocking buffer (PBS containing 1.5% Bovine serum albumin, BSA) for 2 hours at RT. Serum samples were serially diluted in PBS supplemented with 0.05% Tween 20 and 1% BSA and added to the viral protein-or PBS-coated plates for 2 hours at RT. After washing, plates were incubated with horseradish peroxidase (HRP)-conjugated anti-human Ig secondary antibodies diluted in PBS containing 0.05% Tween 20 1% BSA for 45 min at RT. Plates were washed 5 times with PBS-T and developed with TMB substrate reagent set (BD bioscience) with development reaction stopped with 1M H_2_SO_4_. Absorbance was measured at 450 nm on a microplate reader (Infinite 200 PRO, Tecan). To detect RBD-specific IgM, HRP-conjugated anti-human IgM (Southern Biotech) were used at a 1:4000 dilution. To analyze RBD-specific IgG antibody subclasses, HRP-conjugated anti-human IgG1, IgG2, IgG3 and IgG4 (Southern Biotech) were used at a 1:3000 dilutions. To detect SARS-CoV-2-specific IgA1 and IgA2, HRP-conjugated anti-human IgA1 or IgA2 (Southern Biotech) were used at a dilution of 1:2000 and 1:4000, respectively. To quantitate the level of each viral antigen-specific antibody class or subclasses optical density (OD) values were measured and the area under the curve (AUC) derived from optical density measurements of four serial dilutions was determined using Prism 8 (GraphPad). Negative threshold values were set for each immunoglobulin using naïve baseline AUC levels plus 2 times the standard deviations of the mean. Values below the background levels were replaced by the OD value of the blank.

### Data analysis and visualization

GraphPad Prism (version 8.0) was used to conduct statistical analyses. For each experiment, the type of statistical test, summary statistics and levels of significance were specified in the figures and corresponding legends. All tests were performed two-sided with a nominal significance threshold of p < 0.05. For comparisons between time points, paired tests were used.

## Data Availability

Requests for resources and reagents should be directed to and will be fulfilled by the corresponding author, Giuliana Magri.

## Acknowledgments

We would like to thank all the subjects who participated in our studies. This study was supported by the COVID-19 call grant from Generalitat de Catalunya, Department of Health (to G.M and L.D.C.M), grant Miguel Servet research program (to G.M) and by National Health Institute Carlos III (ISCIII) through the project COV20_00508 grant (Co-funded by European Regional Development Fund/European Social Fund “a way to make Europe) (to R.G.).

## Author Contributions

S.T.V. and L.D.C.M performed experiments, analyzed and discussed data and wrote the manuscript; J.M.R, P.D., J.N.B. and C.R.L. recruited study participant and process samples; A.C. participated in data analysis and interpretation and wrote the manuscript; R.G. conceived the study, recruited study participants, discussed data and wrote the manuscript; G.M. conceived the study and designed experiments, analyzed results, discussed data, and wrote the manuscript.

## References

1. Krammer, F. SARS-CoV-2 vaccines in development. Nature vol. 586 516–527 (2020).

2. Polack, F. P. et al. Safety and Efficacy of the BNT162b2 mRNA Covid-19 Vaccine. N. Engl. J. Med. 383, 2603–2615 (2020).

3. Baden, L. R. et al. Efficacy and Safety of the mRNA-1273 SARS-CoV-2 Vaccine. N. Engl. J. Med. 384, 403–416 (2021).

4. Camara, C. et al. Differential effects of the second SARS-CoV-2 mRNA vaccine dose on T cell immunity in naïve and COVID-19 recovered individuals. bioRxiv 2021.03.22.436441 (2021) doi:10.1101/2021.03.22.436441.

5. Widge, A. T. et al. Durability of Responses after SARS-CoV-2 mRNA-1273 Vaccination. N. Engl. J. Med. 384, 80–82 (2021).

6. Goel, R. R. et al. Distinct antibody and memory B cell responses in SARS-CoV-2 naïve and recovered individuals following mRNA vaccination. Sci. Immunol. 6, (2021).

7. Ebinger, J. E. et al. Antibody responses to the BNT162b2 mRNA vaccine in individuals previously infected with SARS-CoV-2. Nat. Med. (2021) doi:10.1038/s41591-021-01325-6.

8. Edara, V. V., Hudson, W. H., Xie, X., Ahmed, R. & Suthar, M. S. Neutralizing Antibodies against SARS-CoV-2 Variants after Infection and Vaccination. JAMA - Journal of the American Medical Association (2021) doi:10.1001/jama.2021.4388.

9. Ju, B. et al. Human neutralizing antibodies elicited by SARS-CoV-2 infection. Nature 584, 115–119 (2020).

10. Suthar, M. S. et al. Rapid Generation of Neutralizing Antibody Responses in COVID-19 Patients. Cell Reports Med. 1, 100040 (2020).

11. Konstantinidis, T. et al. Levels of produced antibodies after vaccination with mRNA vaccine; effect of previous infection with SARS-CoV-2. medRxiv 2021.04.05.21254934 (2021) doi:10.1101/2021.04.05.21254934.

12. Wang, Z. et al. mRNA vaccine-elicited antibodies to SARS-CoV-2 and circulating variants. Nature 592, (2021).

13. Krammer, F. et al. Antibody Responses in Seropositive Persons after a Single Dose of SARS-CoV-2 mRNA Vaccine. N. Engl. J. Med. 384, 1372–1374 (2021).

14. Reynolds, C. J. et al. Prior SARS-CoV-2 infection rescues B and T cell responses to variants after first vaccine dose. Science (2021) doi:10.1126/science.abh1282.

15. Stamatatos, L. et al. mRNA vaccination boosts cross-variant neutralizing antibodies elicited by SARS-CoV-2 infection. Science (80-.). eabg9175 (2021) doi:10.1126/science.abg9175.

16. de Campos-Mata, L. et al. SARS-CoV-2 sculpts the immune system to induce sustained virus-specific naïve-like and memory B cell responses. medRxiv 2021.04.29.21256002 (2021) doi:10.1101/2021.04.29.21256002.

17. Dan, J. M. et al. Immunological memory to SARS-CoV-2 assessed for up to 8 months after infection. Science (80-.). eabf4063 (2021) doi:10.1126/science.abf4063.

18. Wajnberg, A. et al. Robust neutralizing antibodies to SARS-CoV-2 infection persist for months. Science (80-.). 370, 1227–1230 (2020).

19. Hartley, G. E. et al. Rapid generation of durable B cell memory to SARS-CoV-2 spike and nucleocapsid proteins in COVID-19 and convalescence. Sci. Immunol. 5, (2020).

20. Gaebler, C. et al. Evolution of antibody immunity to SARS-CoV-2. Nature 1–6 (2021) doi:10.1038/s41586-021-03207-w.

21. Vidarsson, G., Dekkers, G. & Rispens, T. IgG subclasses and allotypes: From structure to effector functions. Front. Immunol. 5, (2014).

22. Lu, L. L., Suscovich, T. J., Fortune, S. M. & Alter, G. Beyond binding: Antibody effector functions in infectious diseases. Nature Reviews Immunology vol. 18 46–61 (2018).

23. de Campos Mata, L. et al. SARS-CoV-2-specific antibody profiles distinguish patients with moderate from severe COVID-19. medRxiv 2020.12.18.20248461 (2020) doi:10.1101/2020.12.18.20248461.

24. Amanat, F. et al. A serological assay to detect SARS-CoV-2 seroconversion in humans. Nat. Med. 2, (2020).

25. Lederer, K. et al. SARS-CoV-2 mRNA Vaccines Foster Potent Antigen-Specific Germinal Center Responses Associated with Neutralizing Antibody Generation. Immunity 53, 1281-1295.e5 (2020).

26. Chen, K., Magri, G., Grasset, E. K. & Cerutti, A. Rethinking mucosal antibody responses: IgM, IgG and IgD join IgA. Nat. Rev. Immunol. (2020) doi:10.1038/s41577-019-0261-1.

27. Wang, Z. et al. Enhanced SARS-CoV-2 neutralization by dimeric IgA. Sci. Transl. Med. eabf1555 (2020) doi:10.1126/scitranslmed.abf1555.

28. Kim, S. Il et al. Stereotypic neutralizing VHantibodies against SARS-CoV-2 spike protein receptor binding domain in patients with COVID-19 and healthy individuals. Sci. Transl. Med. 13, (2021).

