## Supplementary Materials for "SARS-CoV-2 naïve and recovered individuals show qualitatively different antibody responses following mRNA vaccination"

### Supplementary Material

Fig. S1 (related to Fig. 1 and Fig. 2). mRNA vaccination induces antibody responses irrespective of the sex of the individuals.

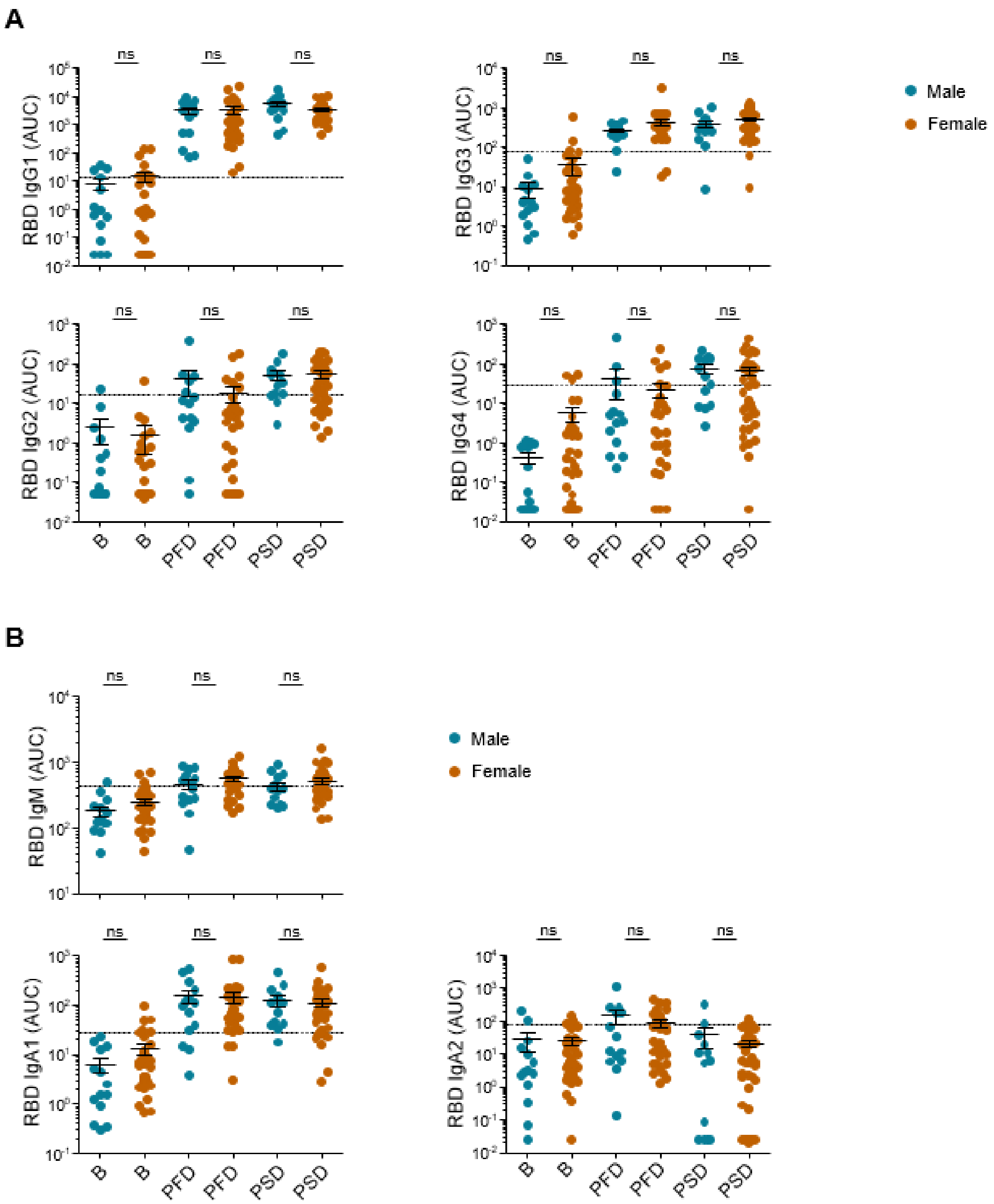


**Fig. S1 (related to Fig. 1 and Fig. 2). mRNA vaccination induces antibody responses irrespective of the sex of the individuals**. (**A**) Area under the curve (AUC) for each of the RBD-specific IgG subclasses analyzed from male and female individuals overtime. (**B**) Area under the curve (AUC) for each of the RBD-specific IgM and IgA subclasses analyzed from male and female individuals overtime. SARS-CoV-2 naïve individuals at baseline were used to establish negative threshold values defined as the naïve AUC mean plus 2 times the standard deviation of the mean. Dashed line indicates negative threshold. Data are presented as individual dots. Two-tailed Mann-Whitney U test was performed to compare antibody responses between male and female. (*P < 0.05, **P < 0.01, and ***P < 0.001). Male, n=14; Female, n=34.
